# Moving Biosurveillance Beyond Coded Data: AI for Symptom Detection from Physician Notes

**DOI:** 10.1101/2023.09.24.23295960

**Authors:** Andrew McMurry, Amy R Zipursky, Alon Geva, Karen L Olson, James Jones, Vlad Ignatov, Timothy Miller, Kenneth D Mandl

**Affiliations:** Computational Health Informatics Program, Boston Children’s Hospital, Boston, Massachusetts, USA; Department of Pediatrics, Harvard Medical School, Boston, Massachusetts, USA; Division of Emergency Medicine, The Hospital for Sick Children, Toronto, Ontario, Canada; Division of Critical Care Medicine, Department of Anesthesiology, Critical Care, and Pain Medicine, Boston Children’s Hospital, Boston, Massachusetts, USA; Department of Anaesthesia, Harvard Medical School, Boston, MA, USA; Department of Biomedical Informatics, Harvard Medical School, Boston, Massachusetts, USA

**Keywords:** Natural language processing, COVID-19, artificial intelligence, public health, biosurveillance, surveillance

## Abstract

**Background:** Real-time surveillance of emerging infectious diseases necessitates a dynamically evolving, computable case definition, which frequently incorporates symptom-related criteria. For symptom detection, both population health monitoring platforms and research initiatives primarily depend on structured data extracted from electronic health records.

**Objective:** To validate and test an artificial intelligence (AI) based Natural Language Processing (NLP) pipeline for detecting COVID-19 symptoms from physician notes.

**Methods:** Subjects in this retrospective cohort study are patients 21 years old and younger, who presented to a pediatric emergency department (ED) at a large academic children’s hospital between March 1, 2020 and May 31, 2022. ED notes for all patients were processed with an NLP pipeline tuned to detect the mention of 11 COVID-19 symptoms based on CDC criteria. For a gold standard, 3 subject matter experts labeled 226 ED notes and had strong agreement (F1=98.6; PPV=97.2; Recall=100.0). F1, PPV, and recall were used to compare the performance of both NLP and ICD-10 to the gold standard chart review. As a formative use case, variations in symptom patterns were measured across SARS-Cov2 variant eras.

**Results:** There were 85,678 ED encounters during the study period, 4.0% with patients with COVID-19. NLP was more accurate at identifying encounters with patients that had any of the COVID-19 symptoms (F1=79.6) than ICD-10 codes (F1=45.1%). NLP accuracy was higher for positive symptoms (recall=93%) than ICD-10 (recall=30%). However, ICD-10 accuracy was higher for negative symptoms (specificity=99.4%) than NLP (specificity=91.7%). Congestion or runny nose showed the highest accuracy difference: NLP F1=82.8%, ICD-10 F1=4.2%. Prevalence of NLP symptoms among patients with COVID-19 differed across variant eras. And patients with COVID-19 were more likely to have each symptom than patients without this disease. Effect sizes (odds ratios) varied across pandemic eras.

**Conclusions:** This study establishes the value of AI based NLP as a highly effective tool for real-time COVID-19 symptom detection in pediatric patients, outperforming traditional ICD-10 methods. It also reveals the evolving nature of symptom prevalence across different virus variants, underscoring the need for dynamic, technology-driven approaches in infectious disease surveillance.

## Introduction

Real time emerging infection surveillance requires a case definition that often involves symptomatology. To detect symptoms, population health monitoring systems and research studies tend to largely rely on structured data from electronic health records (EHRs), including International Classification of Diseases, 10th Revision (ICD-10) coding [1]. However, symptoms are not diagnoses and therefore, may not be consistently coded. We sought to validate and test an open source artificial intelligence (AI) based natural language processing (NLP) pipeline that includes a large language model to detect COVID-19 symptoms from physician notes. As a formative use case, we measured differences in symptom patterns across SARS-CoV2 variant eras.

## Methods

### Study Design and Setting

This is a retrospective cohort study of all patients up to 21 years old presenting to the emergency department (ED) of a large, free-standing, university-affiliated, pediatric hospital between March 1, 2020 and May 31, 2022. The Boston Children’s Hospital Committee on Clinical Investigation found the study to be exempt from human subjects oversight.

### Study Variables

The main dependent variables were a set of 11 COVID-19 symptoms based on Centers for Disease Control and Prevention (CDC) criteria [2]—fever or chills, cough, shortness of breath or difficulty breathing, fatigue, muscle or body aches, headache, new loss of taste or smell, sore throat, congestion or runny nose, nausea or vomiting, and diarrhea. We identified these symptoms by both NLP and ICD-10. For the formative use case, the study period was divided into 3 variant eras defined using Massachusetts COVID-19 data from CoVariant [3]. The pre-Delta era was March 1, 2020 to June 20, 2021, the Delta era was June 21, 2021 to December 19, 2021, and the Omicron era was December 20, 2021 onwards. A diagnosis of COVID-19 was defined as a positive SARS-CoV2 polymerase chain reaction (PCR) test or the presence of ICD-10 code U07.1 for COVID-19 during an encounter.

### AI/NLP Pipeline Development

Three reviewers reached consensus on a symptom concept dictionary [4] to capture each of the 11 COVID-19 symptoms. They relied on the Unified Medical Language System [5] which has a near comprehensive list of symptom descriptors [6] including SNOMED coded clinical terms [7], ICD-10 codes for administrative billing, abbreviations, and common language for patients [8]. The open source and free Apache cTAKES natural language processing pipeline was tuned to recognize and extract coded concepts for positive symptom mentions (based on the dictionary) from physician notes [9]. Apache cTAKES utilizes a NegEx algorithm which can help address negation [9–12]. To further address negation, we incorporated a large language model, BERT, fine-tuned for negation classification on clinical text [13,14].

### Gold Standard

Two reviewers established a gold standard by manually reviewing physician ED notes. After all notes were labeled by the cTAKES pipeline, a sample of 226 ED notes was loaded into Label Studio [15], an open source application for ground truth labeling. These notes were from patients both with and without COVID-19, and were selected to ensure that each of the 11 symptoms was mentioned in at least 30 ED notes. Some notes mentioned more than one symptom. Using an annotation guide (Supplement 1), 2 reviewers, who were masked from the terms identified by the NLP pipeline for note selection, each labeled 113 notes for mention of the 11 COVID-19 symptoms. As per the guide, only symptoms relevant to the present illness were considered positive mentions. Symptoms were not considered positive mentions if stated as past medical history, family history, social history, or an indication for a medication unrelated to the encounter.

### Inter-rater reliability

The F1 score was used to assess consistency in manual chart review. The F1 score is the balance of recall and positive predictive value (PPV) [16]. It was computed by comparing the annotations of each of the 2 initial reviewers to those of a third reviewer, who independently labeled a subset (n=56, 25%) of notes annotated by the other reviewers. The choice of F1 score as the metric for agreement was informed by the observed high frequency of true negative annotations when they were assigned by chance [9,16,17]. Reliability analyses used Python version 3.10.

### AI/NLP and ICD-10 Accuracy

Accuracy measures of true symptom prevalence for each symptom included F1-score, positive predictive value (PPV), recall (sensitivity) and specificity [18,19].

### Formative use case

The impact of pandemic variant era on COVID-19 symptomatology was examined. Descriptive statistics were used to characterize patients presenting to the ED during each pandemic era.

Symptom prevalence amongst ED patients with COVID-19 was assessed in separate analyses for each symptom using Chi-square analyses of 3x2 tables (pandemic era x symptom presence/absence) with alpha set at .05. Post-hoc Chi-square tests were used to compare each pandemic era with all others using a Bonferroni adjusted alpha of .017. To assess the effect of pandemic era, COVID-19 status, and the interaction of these variables on whether or not a patient had each symptom, logistic regression was used in separate analyses for each symptom. Bonferroni adjusted confidence limits were used for post-hoc analyses. If the interaction term was not significant, main effects for COVID-19 and variant era were reported. Data were analyzed using SAS version 9.4 (SAS Institute Inc.).

## Results

### Study population

There were 59,173 unique patients with 85,678 ED encounters during the study period. Characteristics of the entire study cohort and variant-specific cohorts are summarized in Table 1. A patient could appear in the cohort more than once if they had multiple ED encounters.

**Table 1.** Characteristics of patients at emergency department encounters.

|  |  | **Total**<br>(n=85,678) | **Pre-Delta**<br>(n=38,985) | **Delta**<br>(n=24,432) | **Omicron**<br>(n=22,261) |
| --- | --- | --- | --- | --- | --- |

| Age (years), n (%) |  |  |  |  |  |
| --- | --- | --- | --- | --- | --- |
|  | <5 years | 36,835 (43.0) | 15,403 (39.5) | 11,749 (48.1) | 9,683 (43.5) |
|  | >=5 years | 48,843 (57.0) | 23,582 (60.5) | 12,683 (51.9) | 12,578 (56.5) |
| Sex, n (%) |  |  |  |  |  |
|  | Female | 40,250 (47.0) | 18,659 (47.9) | 11,236 (46.0) | 10,355 (46.5) |
|  | Male | 45,428 (53.0) | 20,326 (52.1) | 13,196 (54.0) | 11,906 (53.5) |
| Race, n (%) |  |  |  |  |  |
|  | American Indian | 147 (0.2) | 64 (0.2) | 54 (0.2) | 29 (0.1) |
|  | Asian | 3,244 (3.8) | 1,457 (3.7) | 949 (3.9) | 838 (3.8) |
|  | African American | 13,354 (15.6) | 6,007 (15.4) | 3,943 (16.1) | 3,404 (15.3) |
|  | Pacific Islander | 81 (0.1) | 28 (0.1) | 24 (0.1) | 29 (0.1) |
|  | White | 34,186 (39.9) | 16,990 (43.6) | 9,093 (37.2) | 8,103 (36.4) |
|  | Not Identified | 34,666 (40.4) | 14,439 (37.0) | 10,369 (42.4) | 9,858 (44.2) |
| COVID-19 Classification Method, n (%) |  |  |  |  |  |
|  | COVID-19 Diagnosis | 3,420 (4.0) | 854 (2.2) | 500 (2.0) | 2066 (9.3) |
|  | PCR Positive | 2,167 (2.5) | 518 (1.3) | 294 (1.2) | 1,355 (6.1) |
|  | ICD-10 Code | 3,305 (3.9) | 820 (2.1) | 458 (1.9) | 2,027 (9.1) |

### Inter-rater reliability

High consistency was demonstrated between Reviewer 3, who labeled a subset of notes, and both Reviewer 1 and Reviewer 2, who each labeled half of the notes chosen to establish the gold standard. F1-scores for the 2 reviewers were 98.8% and 98.4%, respectively. PPV was 97.6% and 96.8%, and recall 100% for both.

### AI/NLP ICD-10 Accuracy

As shown in Table 2, the F1 score for NLP was higher and thus more accurate at identifying encounters with patients that had any of the COVID-19 symptoms than ICD-10. NLP also had higher F1 accuracy for each individual symptom. In addition, NLP sensitivity (recall) of true positive symptoms was higher than ICD-10. However, NLP accuracy of true negative symptoms (specificity) was somewhat lower compared to ICD-10.

**Table 2.**
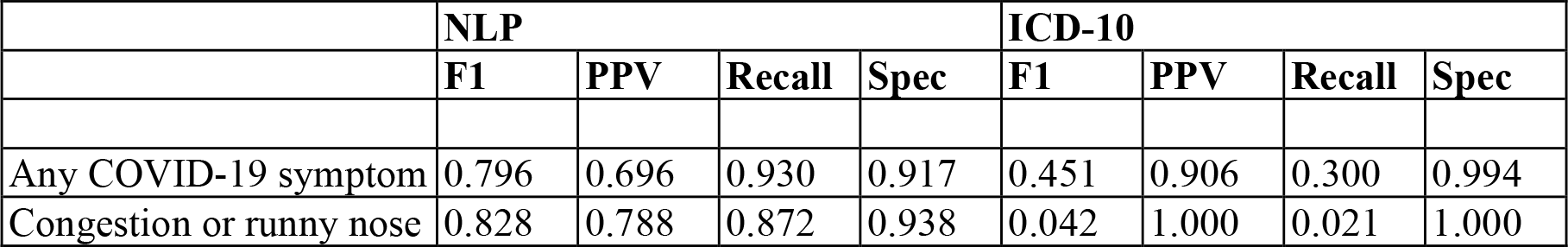
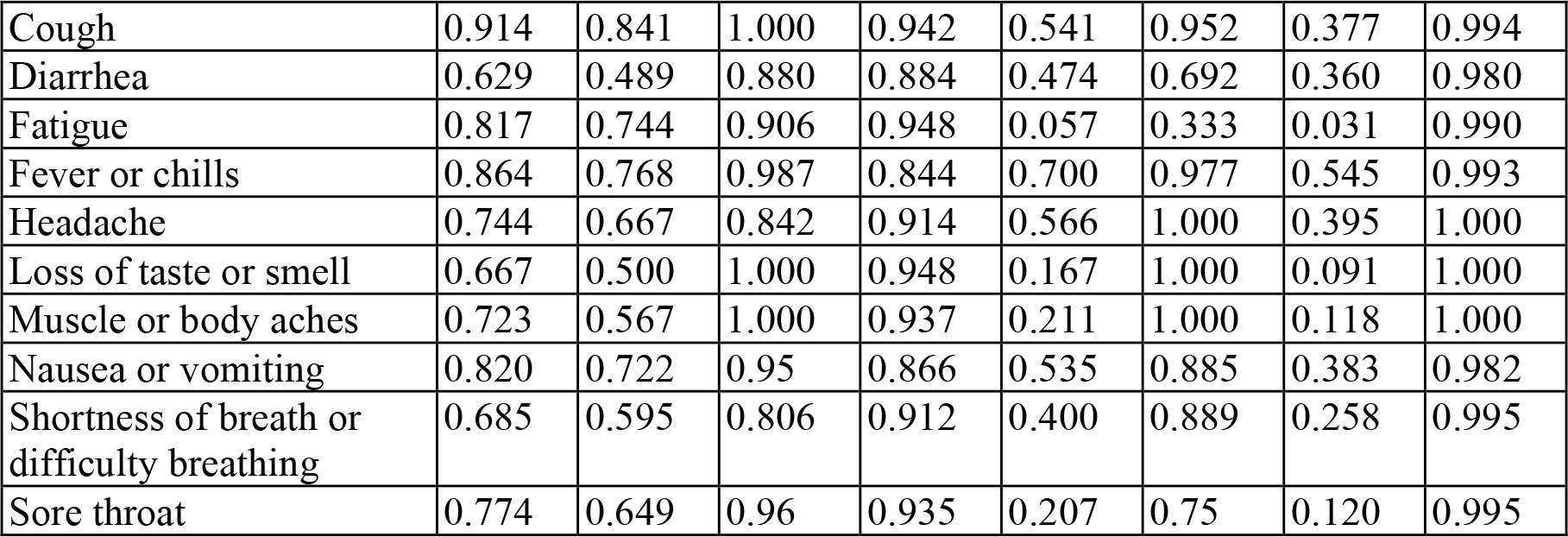
Accuracy of COVID-19 symptom monitoring by NLP and ICD-10. F1: accuracy measure balancing precision and recall, ICD-10: International Classification of Diseases, 10th Revision, NLP: natural language processing, PPV: positive predictive value, Recall: also known as sensitivity, Spec: specificity.

The 2 most prevalent symptoms, cough and fever, had NLP recall scores that were among the highest of the symptoms, and much higher than those for ICD-10 codes. The greatest discrepancy between NLP and ICD-10 F1 accuracy was for congestion or runny nose. The smallest difference was for diarrhea.

### Symptom Prevalence over time

During each month of the study, the percentage of encounters with asymptomatic COVID-19 positive patients was much lower using NLP compared to ICD-10 (Figure 1). Using NLP, the range was from 0 to 19% of encounters (Mean 6, SD 4), while with ICD-10, the range was 22 to 52% (Mean 38, SD 7).

**Figure 1.**
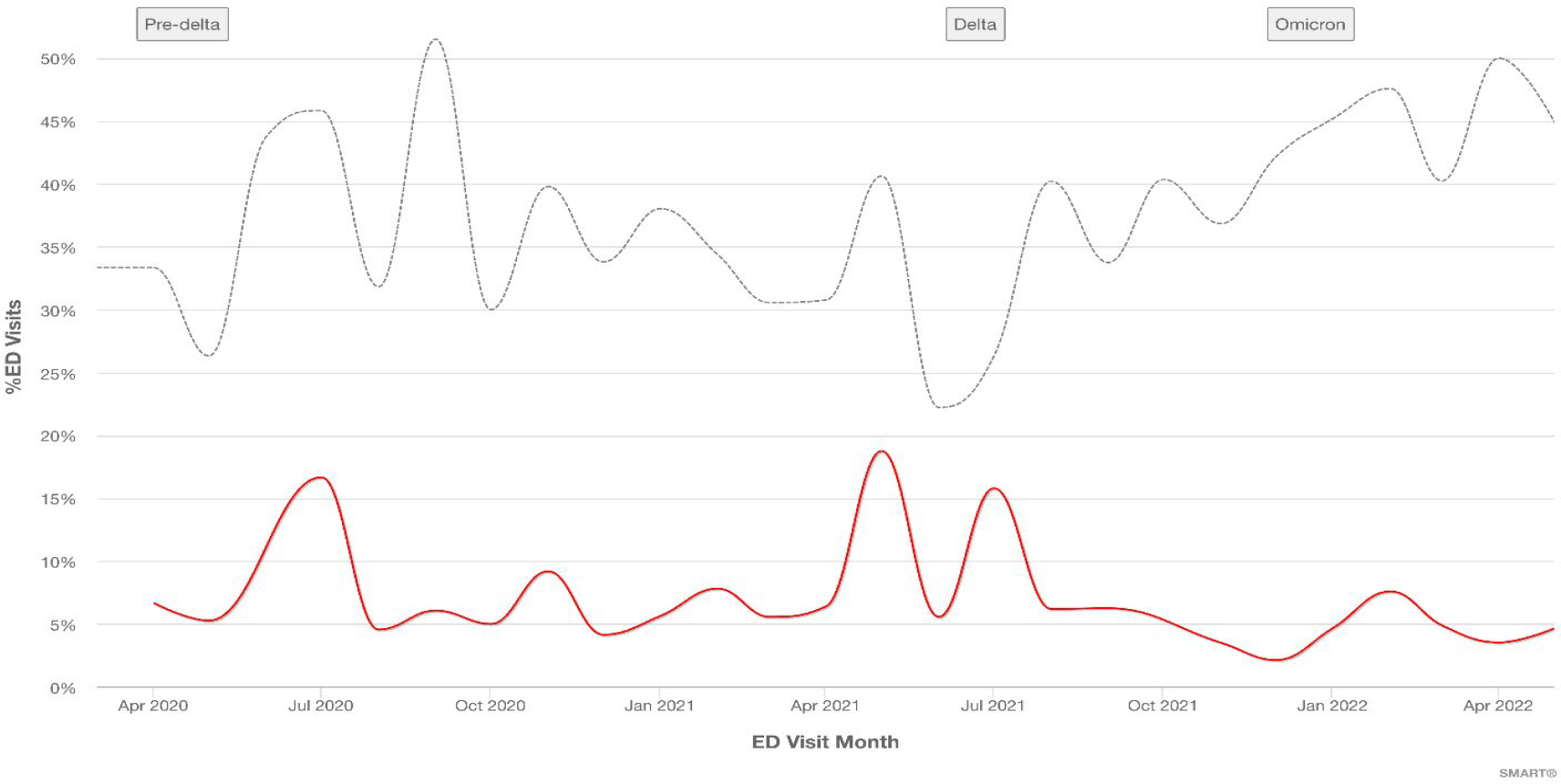
Asymptomatic COVID-19 patients presenting to emergency departments, as measured using NLP and ICD-10. NLP (solid red line), ICD-10 (black dotted line), ED (emergency department).

Monthly prevalence for each symptom was higher using NLP than ICD-10 (Supplement 2). The 2 most prevalent symptoms for encounters with COVID-19 patients, cough and fever, are shown in Figure 2 and Figure 3. On average, cough was identified during 52% (SD 13) of the encounters using NLP, but only 15% (SD 5) using ICD-10. And on average, fever characterized 70% (SD 11) of encounters using NLP, but 41% (SD 9) using ICD-10.

**Figure 2.**
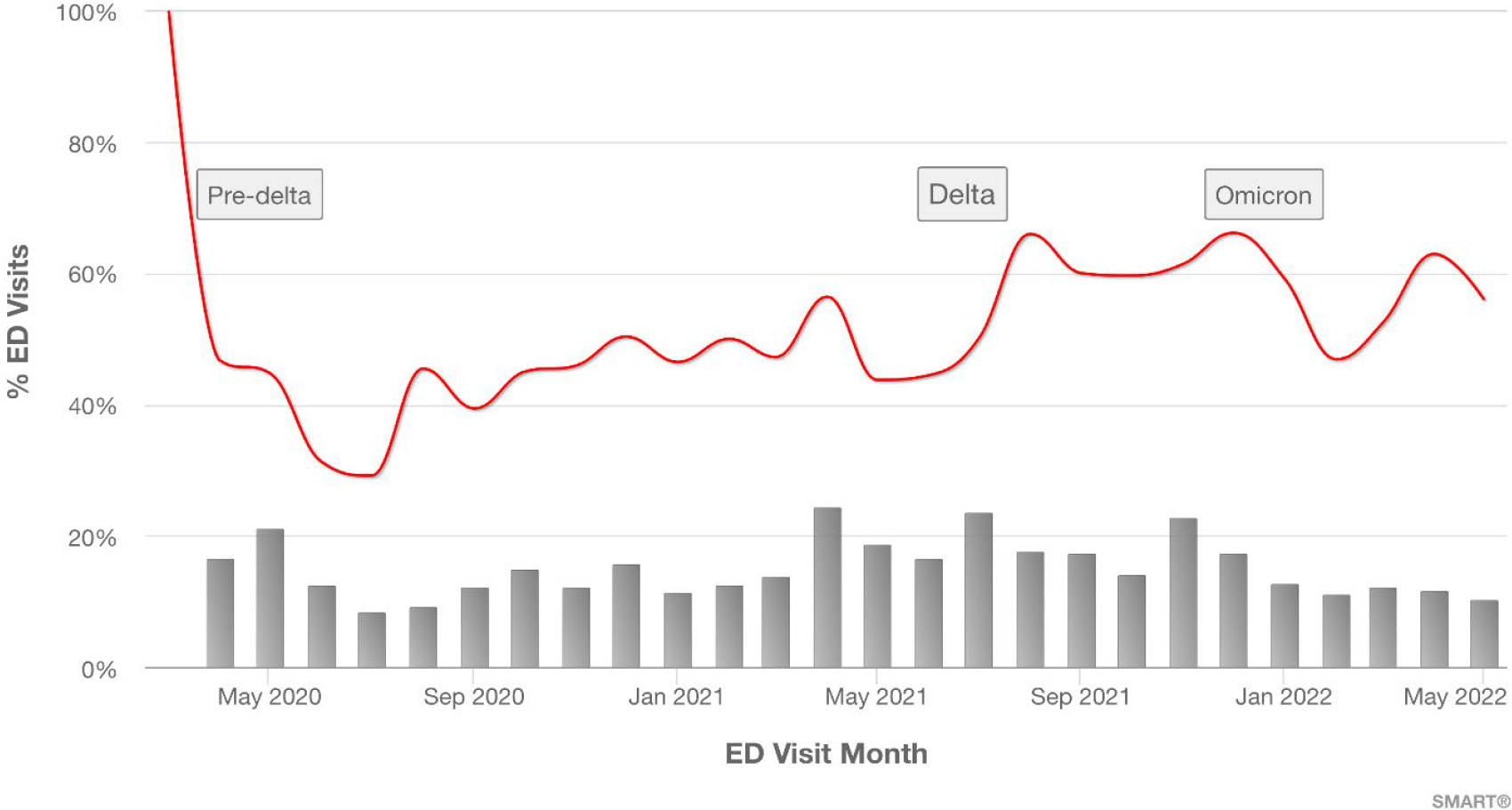
Prevalence of cough during emergency department encounters with patients with COVID-19. NLP (solid red line), ICD-10 (black bars), ED (emergency department).

**Figure 3.**
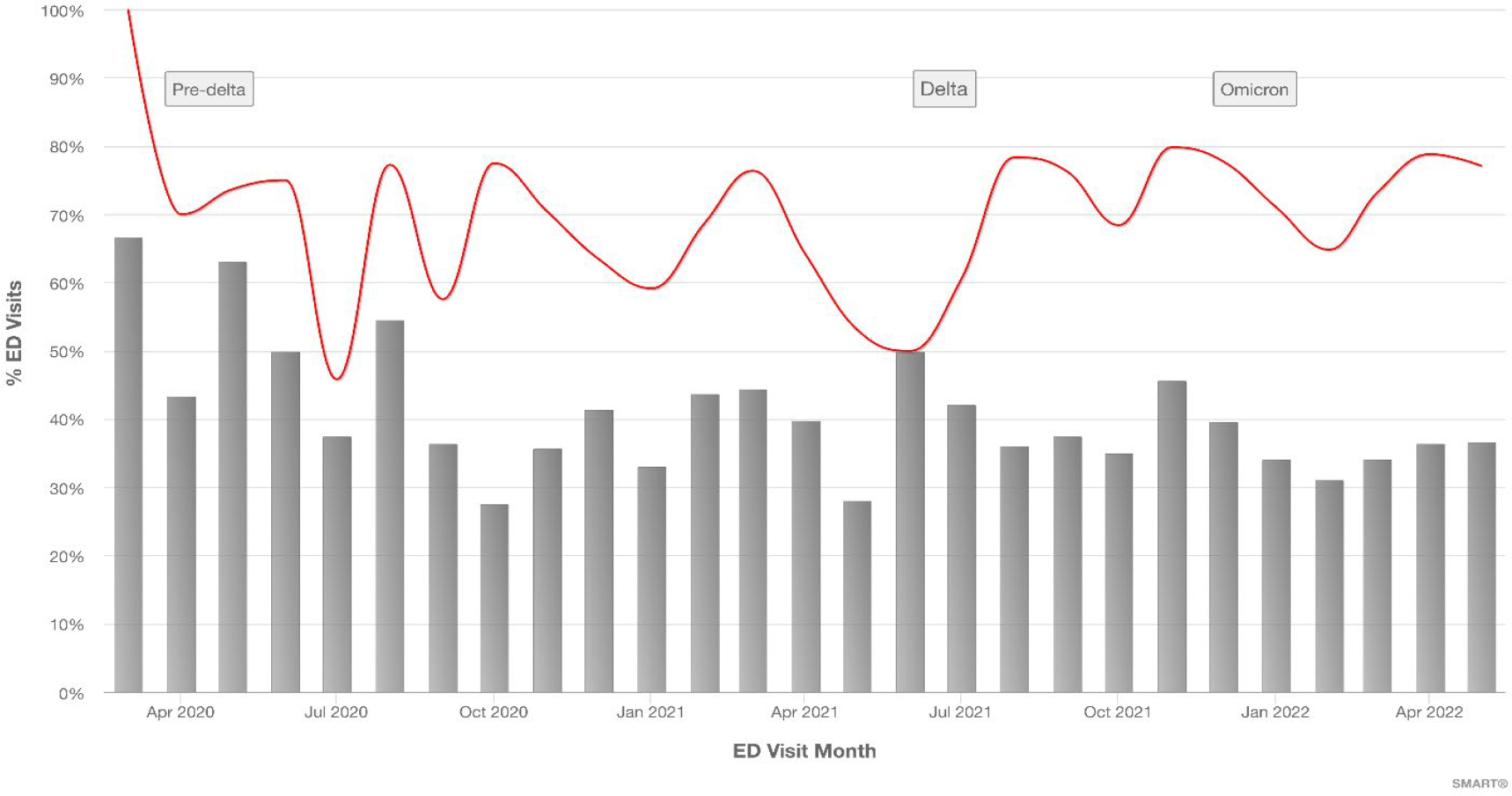
Prevalence of fever during emergency department encounters with patients with COVID-19. NLP (solid red line), ICD-10 (black bars), ED (emergency department).

Using ICD-10, there were many months where individual symptoms were not detected. Of the 27 study months, loss of taste or smell was not detected using ICD-10 during 24 months, nor were muscle or body aches during 13. Three more symptoms had at least 3 consecutive months where each was not detected using ICD-10. These were congestion or runny nose (9), sore throat (8), and fatigue (7). Sporadic months without detection using ICD-10 were observed for headache (5), diarrhea (2), cough (1), and nausea or vomiting (1). Using NLP, sporadic months without detection were observed for just 2 symptoms, loss of taste or smell (6) and sore throat (2).

### Symptom Prevalence across variant eras

Prevalence across variant eras during encounters with patients with COVID-19 differed for each symptom identified by NLP, except for nausea or vomiting and sore throat (Table 3). Post-hoc analyses revealed several patterns. New loss of taste or smell was the only symptom that varied across all 3 eras. It was most prevalent in the pre-Delta era, followed by Delta and then Omicron. Congestion or runny nose, cough, and fever or chills, were more prevalent during Delta and Omicron than during pre-Delta, but Delta did not differ from Omicron. Muscle or body aches were more prevalent during pre-Delta than both Delta and Omicron, but Delta did not differ from Omicron. Diarrhea, fatigue, headache, and shortness of breath were more prevalent during pre-Delta than Omicron but were not different than Delta, and Delta did not differ from Omicron.

**Table 3.** Symptom prevalence by variant era for encounters with patients with COVID-19. Variant eras with the same superscript across a row did not differ in post-hoc analyses.

| Symptom | Pre-Delta (n=854), n(%) | Delta (n=500), n(%) | Omicron (n=2066), n(%) | P value |
| --- | --- | --- | --- | --- |
| Congestion or runny nose | 250 (29.3)<sup>a</sup> | 186 (37.2)<sup>b</sup> | 742 (35.9)<sup>b</sup> | .001 |
| Cough | 402 (47.1)<sup>a</sup> | 309 (61.8)<sup>b</sup> | 1223 (59.2)<sup>b</sup> | <.001 |
| Diarrhea | 188 (22.0)<sup>a</sup> | 92 (18.4)<sup>a,b</sup> | 317 (15.4)<sup>b</sup> | <.001 |
| Fatigue | 129 (15.1)<sup>a</sup> | 72 (14.4)<sup>a,b</sup> | 228 (11.0)<sup>b</sup> | .004 |
| Fever or chills | 561 (65.7)<sup>a</sup> | 376 (75.2)<sup>b</sup> | 1525 (73.8)<sup>b</sup> | <.001 |
| Headache | 185 (21.7)<sup>a</sup> | 92 (18.4)<sup>a,b</sup> | 301 (14.6)<sup>b</sup> | <.001 |
| Muscle or body aches | 110 (12.9)<sup>a</sup> | 39 (7.8)<sup>b</sup> | 164 (7.9)<sup>b</sup> | <.001 |
| Nausea or vomiting | 297 (34.8) | 170 (34.0) | 709 (34.3) | .954 |
| New loss of taste or smell | 57 (6.7)<sup>a</sup> | 9 (1.8)<sup>b</sup> | 9 (0.4)<sup>c</sup> | <.001 |
| Shortness of breath or difficulty breathing | 182 (21.3)<sup>a</sup> | 84 (16.8)<sup>a,b</sup> | 311 (15.1)<sup>b</sup> | <.001 |
| Sore throat | 125 (14.6) | 83 (16.6) | 319 (15.4) | .626 |

<sup>a,b,c</sup> Variant eras with the same superscript across a row did not differ in post-hoc analyses.

Nausea or vomiting and sore throat did not differ by variant era. Chi-square results are in Supplement 3.

### Symptoms by COVID-19 status and variant era

The interaction of COVID-19 status and variant era on the presence of each symptom is shown in Table 4. However, because the interaction was not significant for 2 symptoms, fever and chills and sore throat, main effects for COVID-19 status are shown for both (*P* <.001). The odds ratios indicate that patients with each of these symptoms were more likely to have COVID-19 at an encounter than not have COVID-19. These symptoms were also more likely to occur during Delta and Omicron than during pre-Delta. For the remaining symptoms, the interaction term was significant and odds ratios in each variant era are shown in the table. The odds ratios comparing patients with COVID-19 to those without the disease differed among the variant eras. Several patterns were observed. For congestion or runny nose, cough, fatigue, headache, muscle or body aches, new loss of taste or smell, shortness of breath or difficulty breathing, each symptom was more likely to be observed in patients with COVID-19. However, effect sizes (odds ratios) differed among pandemic eras. For diarrhea, this symptom was more likely for COVID-19 patients in the pre-Delta and Delta eras, but not during Omicron. And nausea was more likely only in the pre-Delta era. Significant odds ratios ranged in size from 1.3 to 26.7 (Mean 4.6). The logistic regression results are in Supplement 4.

**Table 4.** Effect of COVID-19 status and variant era on the presence of each symptom. Odds ratios compare patients with COVID-19 at an ED encounter to patients without the disease. If the interaction term was significant, the effect of COVID-19 during each variant era is shown. Otherwise, the effect for COVID-19 is shown.

| Symptom | Pandemic variant era | Odds Ratio (95% CL<sup>a</sup>) | Interaction *P* value<sup>b</sup> |
| --- | --- | --- | --- |
| Congestion or runny nose |  |  |  |
|  | Pre-Delta | 3.62 (3.11-4.21) | <.001 |
|  | Delta | 2.27 (1.89-2.72) |  |
|  | Omicron | 2.46 (2.23-2.71) |  |
| Cough |  |  |  |
|  | Pre-Delta | 4.84 (4.22-5.55) | <.001 |
|  | Delta | 3.64 (3.03-4.37) |  |
|  | Omicron | 3.54 (3.23-3.88) |  |
| Diarrhea |  |  |  |
|  | Pre-Delta | 2.23 (1.89-2.63) | <.001 |
|  | Delta | 1.42 (1.13-1.79) |  |
|  | Omicron | 1.05 (0.92-1.19) |  |
| Fatigue |  |  |  |
|  | Pre-Delta | 3.22 (2.65-3.90) | 0.010 |
|  | Delta | 3.42 (2.64-4.42) |  |
|  | Omicron | 2.36 (2.03-2.75) |  |
| Fever or chills |  | 4.82 (4.46-5.21) | 0.662 |
| Headache |  |  |  |
|  | Pre-Delta | 2.33 (1.98-2.76) | <.001 |
|  | Delta | 2.09 (1.66-2.63) |  |
|  | Omicron | 1.52 (1.33-1.73) |  |
| Muscle or body aches |  |  |  |
|  | Pre-Delta | 5.96 (4.83-7.36) | 0.006 |
|  | Delta | 4.75 (3.38-6.67) |  |
|  | Omicron | 3.78 (3.14-4.55) |  |
| Nausea or vomiting |  |  |  |
|  | Pre-Delta | 1.30 (1.13-1.50) | 0.006 |
|  | Delta | 1.03 (0.86-1.25) |  |
|  | Omicron | 0.98 (0.89-1.08) |  |
| New loss of taste or smell |  |  |  |
|  | Pre-Delta | 26.66 (19.13-37.14) | 0.049 |
|  | Delta | 11.83 (5.68-24.65) |  |
|  | Omicron | 11.04 (4.25-28.64) |  |
| Shortness of breath or difficulty breathing |  |  |  |
|  | Pre-Delta | 2.62 (2.22-3.10) | <.001 |
|  | Delta | 1.70 (1.34-2.16) |  |
|  | Omicron | 1.57 (1.38-1.79) |  |
| Sore throat |  | 2.45 (2.22-2.70) | 0.274 |

<sup>a</sup>CL: Bonferroni adjusted confidence limits in post-hoc analyses.
<sup>b</sup>Type 3 test of the interaction term (variant era x COVID-19) in a logistic regression analysis.

## Discussion

### Principal Findings

We find evidence that AI-based NLP of physician notes is a superior method for capturing patient symptoms for real-time biosurveillance than reliance on traditional approaches using ICD-10. NLP was more sensitive than ICD-10 codes in identifying symptoms and some symptoms could only be detected using NLP. As a form of internal validation, the symptoms identified by the CDC as associated with COVID-19 were more prevalent in patients with than without this disease.

### Comparison with Prior Work

The study was also able to capture a nuanced picture of symptom prevalence and odds across different SARS-CoV-2 variant eras. Consistent with previous literature, symptom patterns changed over time as new variants emerged. Variants may present with differences in symptomatology as a result of a number of factors including differences in mutations in spike proteins, receptor binding, and ability to escape host antibodies [20]. As has been previously reported [21–25], we found that fever or chills was the most common COVID-19 symptom across variants. In our cohort, shortness of breath was less common in the Omicron compared to pre-Delta era. Omicron has less of an ability to replicate in the lungs compared to the bronchi, which may explain why this symptom became less common [26]. Studies have reported sore throat as a common symptom in the Omicron era, but we did not observe a significant difference across eras [27,28]. It is possible that we did not see a higher prevalence of sore throat in the Omicron era because it may be more challenging for pediatric patients to describe this symptom. One study found that sore throat was observed more often in those 5-20 years old compared to those 0-4 [28]. Similarly, a study reported that sore throat was more common in those greater than or equal to 13 years old in Omicron compared to Delta [29]. In our study cohort, approximately half of the patients were less than 5 years old. As children this age may not be able to describe their symptoms well, symptoms that are also signs, like fever or cough, might be more commonly documented in physician notes than symptoms like sore throat. New loss of taste or smell was most prevalent in the pre-Delta era, followed by Delta and then Omicron in this study. This symptom has been reported less commonly in Omicron [27,28]. Studies have postulated that patients with Omicron are less likely to present with loss of taste or smell as this variant has less penetration of the mucus layer and therefore may be less likely to infect the olfactory epithelium [30].

### Limitations

There were important limitations in our use of NLP. The NLP pipeline does not account for vital signs and so fever may not have been detected with the pipeline if it was documented in a patient’s vital signs rather than the clinical text. The cTAKES tool in the pipeline lacks the temporal context to ascertain if the mention of a symptom in a note is a new symptom or a prior symptom. We modified our technique because of this, but nevertheless may have overestimated the prevalence of symptoms in our study. Future work will involve filtering by note section so that certain components of a note like past medical history are not included. Finally, we utilized two techniques to recognize negation, but some negated symptoms (e.g., “patient had no cough”) were still captured as positive symptom mentions leading to possible overestimation of symptom prevalence.

Our formative study had some limitations. First, we examined COVID-19 symptoms in patients presenting to a single urban pediatric ED. Patients presenting to outpatient settings, who likely had milder symptoms, were not included and our results may reflect patients with more severe symptoms. And because the setting was a single site, results may not generalize to other EDs.

Second, we defined COVID-19 status as positive if a patient had a PCR positive test for COVID-19 or an appropriate ICD-10 code at the ED encounter. Patients who were COVID-19 positive on a test at home or at an outside center may not have been captured by this definition even if they presented to the ED with COVID-19 [31]. Additionally, symptoms may have differed across variant eras as a result of COVID-19 vaccinations or previous infections rather than variant differences. Literature in adults shows that vaccination is associated with a decrease in systemic symptoms [32]. The United States Food and Drug Administration authorized the use of the COVID-19 vaccine in October of 2021, during the Delta era and prior to the Omicron era, for children 5 to 11 years old [33]. Vaccination rates for pediatric patients vary by age group in Massachusetts; of those 0-19 years of age, 3%-57% have received a primary series but have not been boosted, and 3%-18% have been boosted since September 1, 2022 [34]. As such, some patients in the Delta and Omicron eras may have been vaccinated or had previous COVID-19 infections [35].

## Conclusions

In an era where rapid and accurate infectious disease surveillance is crucial, this study underscores the transformative potential of AI-based NLP for real-time symptom detection, significantly outperforming traditional methods like ICD-10 coding. The dynamic adaptability of NLP technology allows for the nuanced capture of evolving symptomatology across different virus variants, offering a more responsive and precise toolkit for biosurveillance efforts. Its integration into existing healthcare infrastructure could be a game-changer, elevating our capabilities to monitor, understand, and ultimately control the spread of emerging infectious diseases.

## Data Availability

All data produced in the present study are available upon reasonable request to the authors.

## Acknowledgements

Conceptualized by KDM, AM, TM. Software and Analysis by KLO, AM, ARZ, JJ, AG, VI. First draft of manuscript by AM, KDM, ARZ. Manuscript edits by KLO, AG. Funding obtained by KDM. Work supported by the Centers for Disease Control and Prevention of the U.S. Department of Health and Human Services (HHS) as part of a financial assistance award (KDM, AM, DG, TM, KLO, IG) The contents are those of the author(s) and do not necessarily represent the official views of, nor an endorsement, by CDC/HHS, or the U.S. Government. ARZ was supported by a training grant from the National Institute of Child Health and Human Development, T32HD040128.

## Conflicts of Interest

None Declared.

### Abbreviations

AI: artificial intelligence
ED: emergency department
EHR: electronic health record
ICD-10: International Classification of Diseases, 10th Revision
NLP: natural language processing
PPV: positive predictive value

## Supplements

### Supplement 1

COVID-19 symptoms annotation guide.

### Supplement 2

Detection of COVID-19 symptoms using NLP and ICD-10 by month for ED encounters with COVID-19 positive patients.

### Supplement 3

Chi-square analysis of COVID-19 symptom prevalence by pandemic variant era for ED encounters with COVID-19 positive patients. Symptoms were detected using NLP.

### Supplement 4

Logistic regression analysis of the effect of COVID-19 status, pandemic variant era, and their interaction on symptom status for ED encounters. Symptoms were detected using NLP.

