## Supplement 1 for "Moving Biosurveillance Beyond Coded Data: AI for Symptom Detection from Physician Notes"

**COVID-19 Symptom Annotation Guide**

Three expert reviewers reached consensus on three criteria for annotating positive mentions of COVID-19 symptoms in ED physician notes. The three criteria are applied in series (1) encounter time criteria; (2) ED note section criteria; and (3) symptom specific criteria.

**Criteria 1: Encounter time.** Symptoms must be relevant to the present ED encounter either as the reason for visit, documented symptom, or observed sign.

| Encounter Time | Include | Exclude |
|---|---|---|
| Prior to encounter | Chief complaint or patient reason for visit | Symptoms recorded for an illness or condition unrelated to the current encounter |
| Present ED encounter | Yes | Symptoms recorded for an illness or condition unrelated to the current encounter |
| Future encounter | Never | Always |

**Criteria 2: ED Note Section.** Patient symptoms are present and not related to past medical history or a medication prescribed unrelated to the present encounter.

| ED Note Section | Include | Exclude |
|---|---|---|
| Chief complaint | Symptom present | |
| History of presenting illness | Symptom present | |
| Review of systems | Symptom present | |
| Physical exam | Symptom present | |
| Vital signs | Fever present | |
| Family history | | Always (e.g. brother had a cough) |
| Past medical history | | Always (e.g. patient had cough three months ago) |
| Social history | | Always (e.g. contacts in daycare have vomiting) |
| Medication list | | Always (e.g. albuterol PRN for cough) |
| Investigations | | Always (e.g. opacity on chest x-ray) |
| Assessment and plan, course, evaluation | Symptom present | |
| Final diagnosis | Symptom present | |
| Discharge Instructions | Symptom present | |

**Criteria 3: Symptom specific.** Positive symptom mentions explicitly state the symptom or a predefined synonym.

| Symptom specific | Include | Exclude |
|---|---|---|
| New loss of taste or smell | Anosmia, loss of taste, loss of smell | Injury related to loss of taste/smell |
| Congestion or runny nose | Rhinorrhea, congestion, discharge, nose is dripping, running, or stuffy | |
| Cough | Tussive or post-tussive, cough is unproductive, productive, dry, wet, or producing sputum | Wheeze, crackles, croup |
| Diarrhea | Diarrhea or watery stool | Loose stool, bloody stool |
| Fatigue | Fatigue, tired, exhausted, weary, malaise, feeling generally unwell | Looked ill |
| Fever or chills | Fever, pyrexia, chills, or temperature >= 100.4 °F [38 °C] | Afebrile, felt warm |
| Headache | HA/headache, migraine, cephalgia, head pain | Headache due to injury |
| Muscle or body aches | Myalgias, myoneuralgia, muscle or body aches, soreness, generalized aches and pains | Localized pain, injury, abdominal pain, lower back pain |
| Nausea or vomiting | Nausea, vomiting, emesis, throw up, queasy, regurgitated | Gastritis, gastroparesis |
| Shortness of breath or difficulty breathing | Dyspnea, breathing is short, difficult, increased, labored, or distressed | BiPAP, CPAP, oxygen need |
| Sore throat | Sore throat, throat pain, pharyngeal pain, pharyngitis, odynophagia | Streptococcus, dysphagia, hoarseness, red throat |
