## Supplement 2 for "Moving Biosurveillance Beyond Coded Data: AI for Symptom Detection from Physician Notes"

**Supplement 2. Time series figures**. Prevalence of 11 COVID-19 symptoms detected by natural language processing and ICD-10 coding during emergency department encounters with patients with COVID-19.

Supplementary information for:

Moving Biosurveillance Beyond Coded Data: AI for Symptom Detection from Physician Notes

Andrew McMurry,[1,2] Amy R Zipursky,[1,3] Alon Geva,[1,4,5] Karen L Olson,[1,2] James Jones,[1,2] Vlad Ignatov,[1] Timothy Miller,[1,2] Kenneth D Mandl[1,2,6,*]

[1] Computational Health Informatics Program, Boston Children's Hospital, Boston, Massachusetts, United States of America
[2] Department of Pediatrics, Harvard Medical School, Boston, Massachusetts, United States of America
[3] Division of Emergency Medicine, The Hospital for Sick Children, Toronto, Ontario, Canada
[4] Division of Critical Care Medicine, Department of Anesthesiology, Critical Care, and Pain Medicine, Boston Children's Hospital, Boston, Massachusetts, United States of America
[5] Department of Anaesthesia, Harvard Medical School, Boston, MA, USA
[6] Department of Biomedical Informatics, Harvard Medical School, Boston, Massachusetts, United States of America

*Corresponding author
 (KDM)

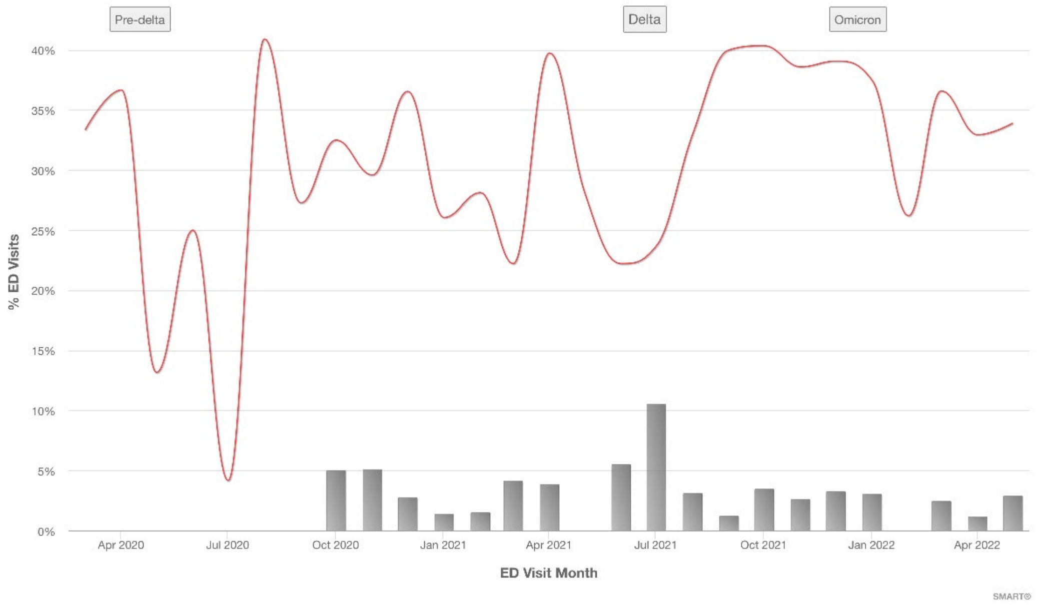


**Figure 1. Prevalence of congestion during emergency department encounters with patients with COVID-19**. NLP (solid red line): natural language processing, ICD-10 (black bars): International Classification of Diseases, 10th Revision, ED: emergency department.

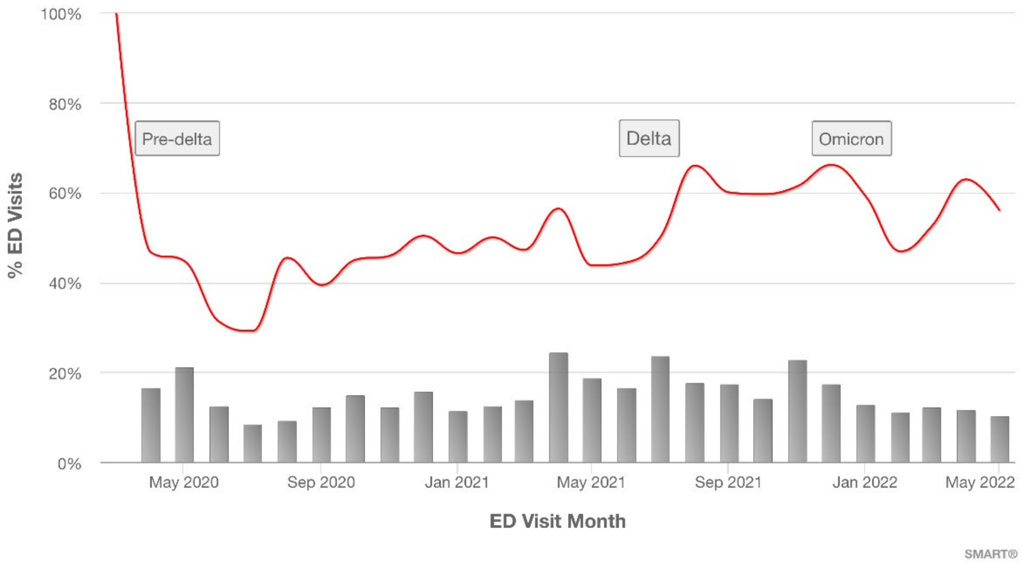


**Figure 2. Prevalence of cough during emergency department encounters with patients with COVID-19**. NLP (solid red line): natural language processing, ICD-10 (black bars): International Classification of Diseases, 10th Revision, ED: emergency department.

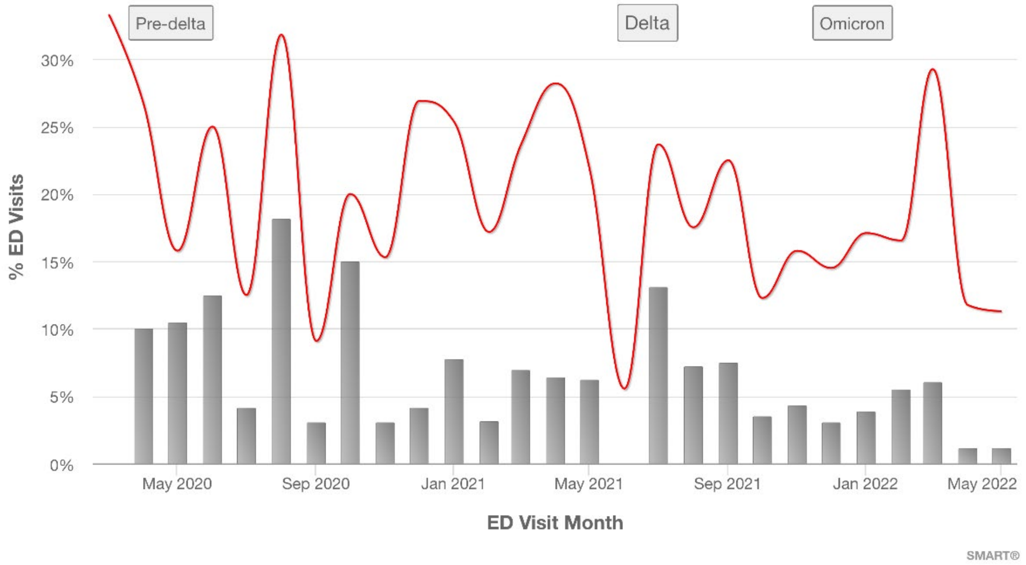


**Figure 3. Prevalence of diarrhea during emergency department encounters with patients with COVID-19**. NLP (solid red line): natural language processing, ICD-10 (black bars): International Classification of Diseases, 10th Revision, ED: emergency department.

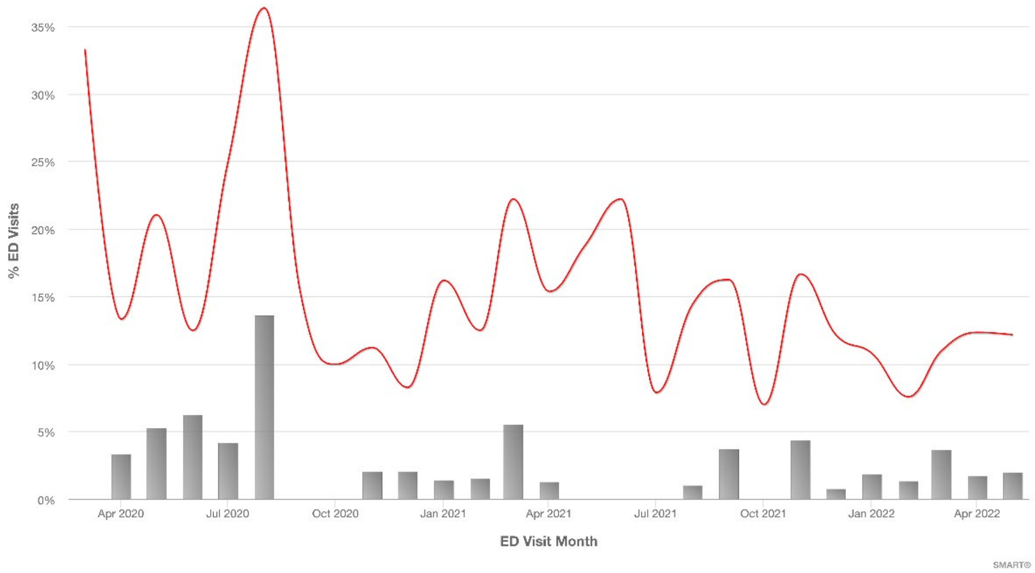


**Figure 4. Prevalence of fatigue during emergency department encounters with patients with COVID-19**. NLP (solid red line): natural language processing, ICD-10 (black bars): International Classification of Diseases, 10th Revision, ED: emergency department.

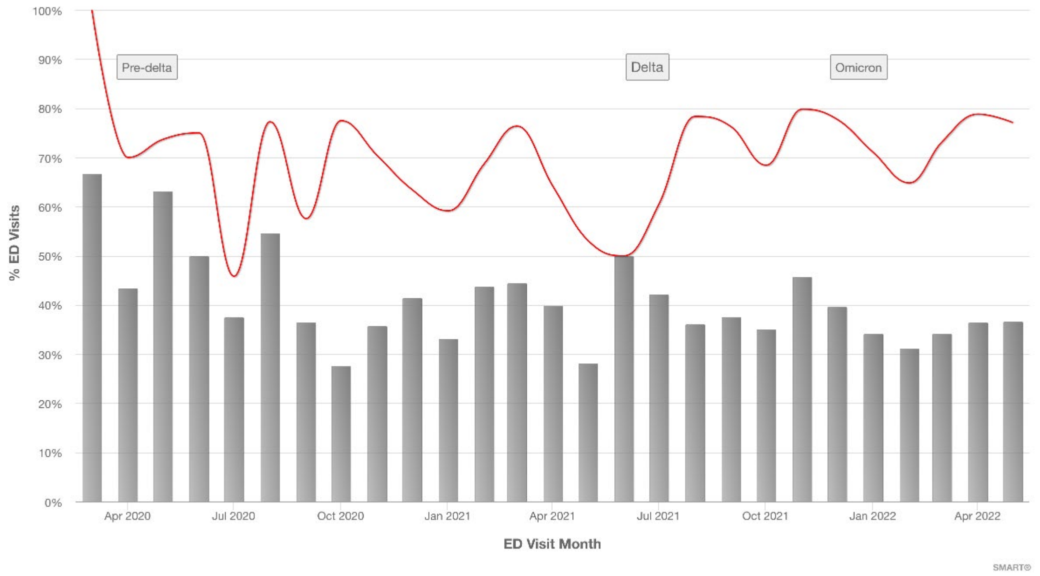


**Figure 5. Prevalence of fever during emergency department encounters with patients with COVID-19**. NLP (solid red line): natural language processing, ICD-10 (black bars): International Classification of Diseases, 10th Revision, ED: emergency department.

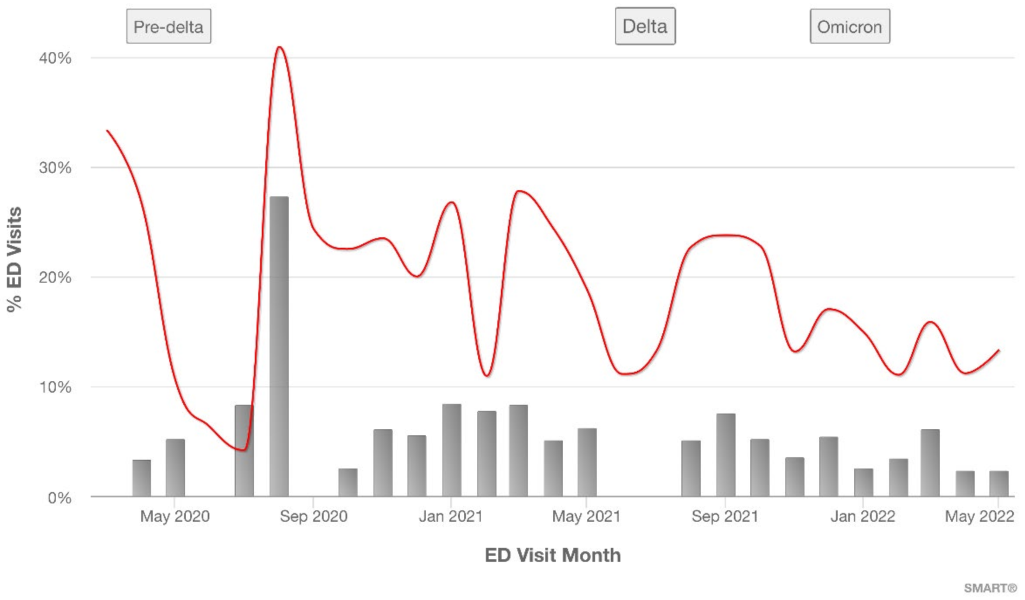


**Figure 6. Prevalence of headache during emergency department encounters with patients with COVID-19**. NLP (solid red line): natural language processing, ICD-10 (black bars): International Classification of Diseases, 10th Revision, ED: emergency department.

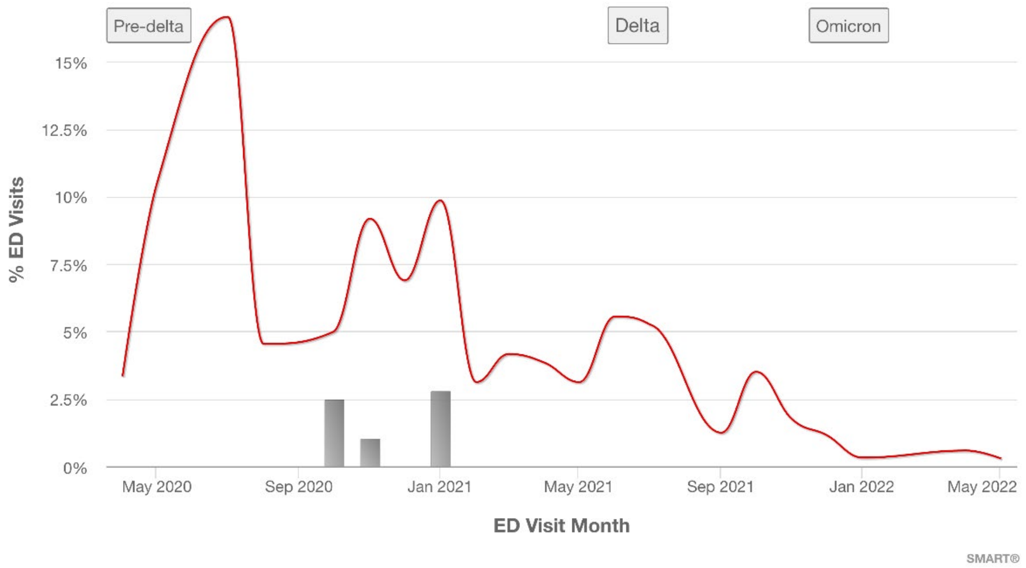


**Figure 7. Prevalence of loss of taste or smell during emergency department encounters with patients with COVID-19**. NLP (solid red line): natural language processing, ICD-10 (black bars): International Classification of Diseases, 10th Revision, ED: emergency department.

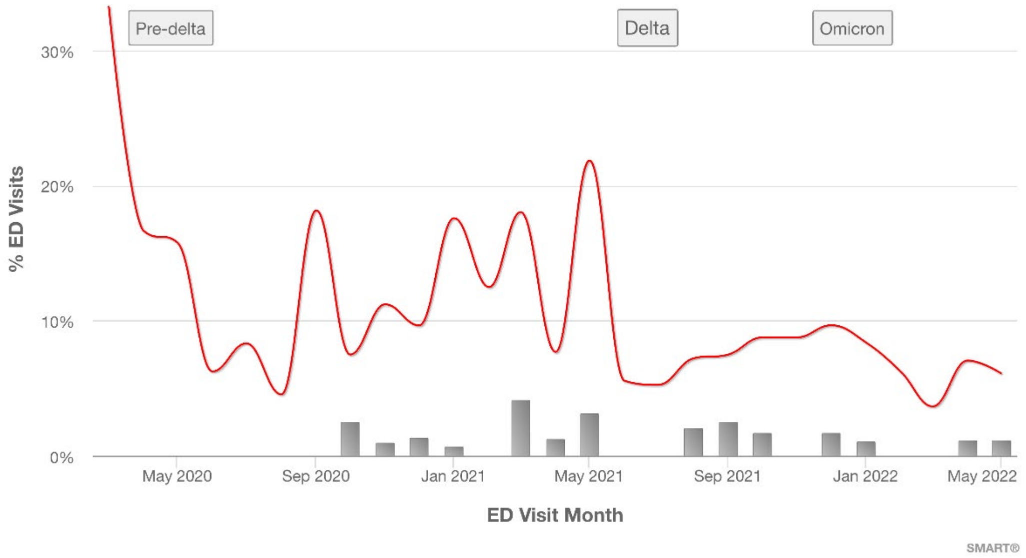


**Figure 8. Prevalence of muscle or body aches during emergency department encounters with patients with COVID-19**. NLP (solid red line): natural language processing, ICD-10 (black bars): International Classification of Diseases, 10th Revision, ED: emergency department.

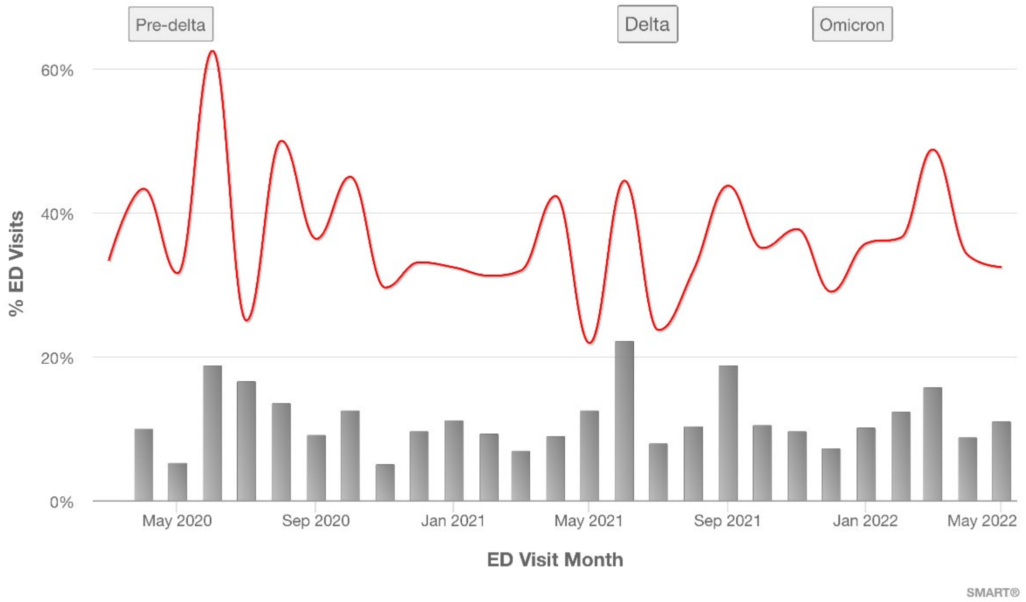


**Figure 9. Prevalence of nausea during emergency department encounters with patients with COVID-19**. NLP (solid red line): natural language processing, ICD-10 (black bars): International Classification of Diseases, 10th Revision, ED: emergency department.

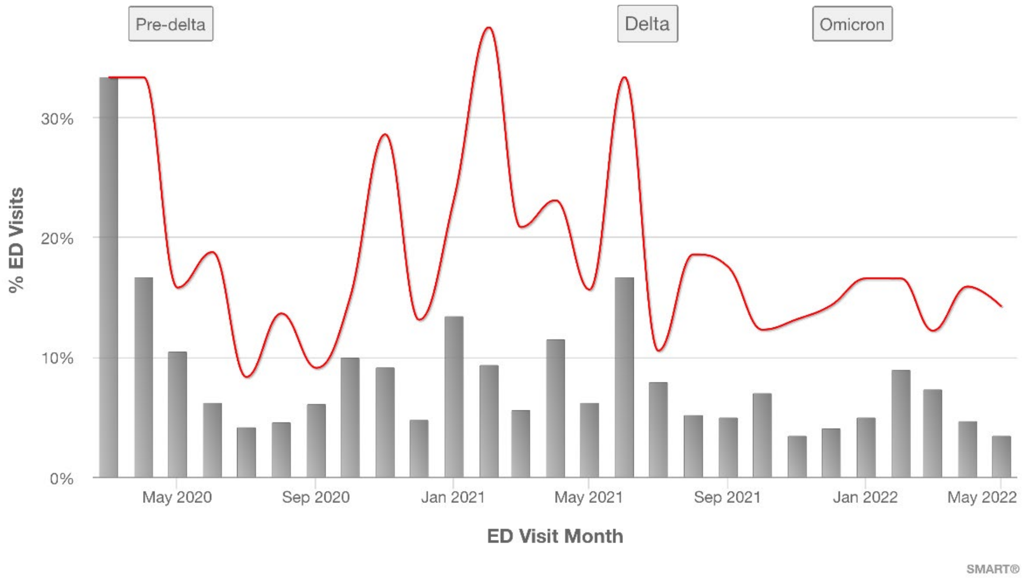


**Figure 10. Prevalence of shortness of breath or difficulty breathing during emergency department encounters with patients with COVID-19**. NLP (solid red line): natural language processing, ICD-10 (black bars): International Classification of Diseases, 10th Revision, ED: emergency department.

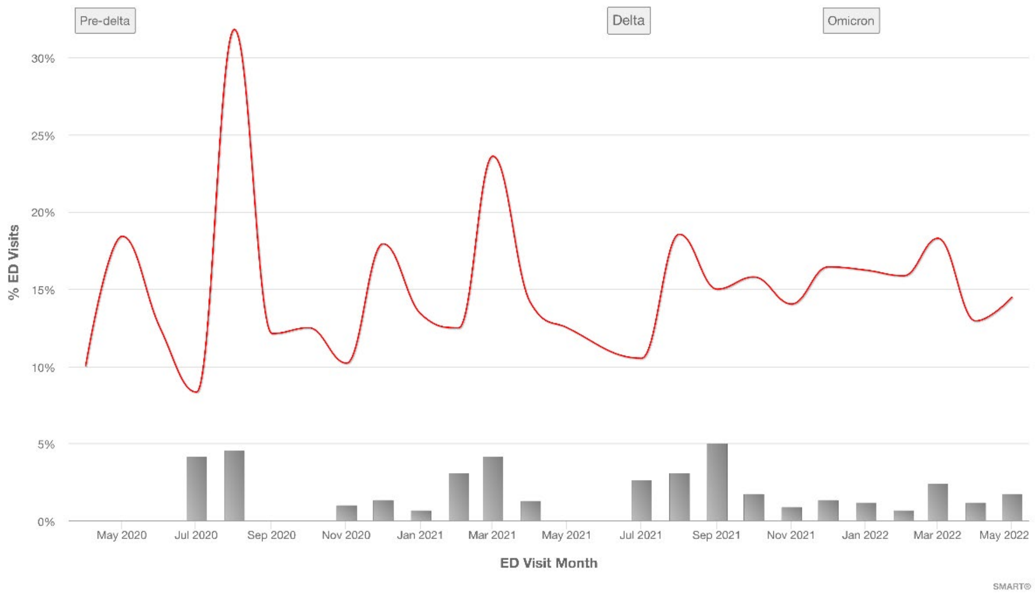


**Figure 11. Prevalence of sore throat during emergency department encounters with patients with COVID-19**. NLP (solid red line): natural language processing, ICD-10 (black bars): International Classification of Diseases, 10th Revision, ED: emergency department.
